# Proposed Clinical Indicators for Efficient Screening and Testing for COVID-19 Infection from Classification and Regression Trees (CART) Analysis

**DOI:** 10.1101/2020.05.11.20097980

**Authors:** Richard K. Zimmerman, Mary Patricia Nowalk, Todd Bear, Rachel Taber, Theresa M. Sax, Heather Eng, GK. Balasubramani

**Author notes:** **Address correspondence to:** Richard K. Zimmerman, MD, Suite 520 Schenley Place, 4420 Bayard Street, Pittsburgh PA 15260. Telephone: 412/383-2354;.

## Abstract

**Background:** The introduction and rapid transmission of SARS CoV2 in the United States resulted in implementation of methods to assess, mitigate and contain the resulting COVID-19 disease based on limited knowledge. Screening for testing has been based on symptoms typically observed in inpatients, yet outpatient symptom complexes may differ.

**Methods:** Classification and regression trees (CART) recursive partitioning created a decision tree classifying enrollees into laboratory-confirmed cases and non-cases. Demographic and symptom data from patients ages 18-87 years who were enrolled from March 29-April 26, 2020 were included. Presence or absence of SARSCoV2 was the target variable.

**Results:** Of 736 tested, 55 were positive for SARS-CoV2. Cases significantly more often reported chills, loss of taste/smell, diarrhea, fever, nausea/vomiting and contact with a COVID-19 case, but less frequently reported shortness of breath and sore throat. A 7-terminal node tree with a sensitivity of 96% and specificity of 53%, and an AUC of 78% was developed. The positive predictive value for this tree was 14% while the negative predictive value was 99%. Almost half (44%) of the participants could be ruled out as likely non-cases *without* testing.

**Discussion:** Among those referred for testing, negative responses to three questions could classify about half of tested persons with low risk for SARS-CoV2 and would save limited testing resources. These questions are: was the patient in contact with a COVID-19 case? Has the patient experienced 1) a loss of taste or smell; or 2) nausea or vomiting? The outpatient symptoms of COVID-19 appear to be broader than the well-known inpatient syndrome.

## Introduction

The introduction of the SARS CoV2 virus into the United States was earlier than anticipated. Furthermore, the rapid transmission of SARS CoV2 resulted in implementation of methods to assess, mitigate and contain the resulting COVID-19 disease that were based on limited knowledge of its epidemiology. Given potentially insufficient testing supplies, but the need to rapidly identify cases, access to testing was based on symptoms typical of an acute respiratory infection, such as cough, fever, and shortness of breath, as has been reported to be emblematic of COVID-19 infection.^1^ A previous report noted that use of cough, fever, shortness of breath and sore throat as the sole screening criteria for COVID-19 infection among health care personnel might have missed 17% of symptomatic cases.^2^ Gastrointestinal and non-acute respiratory illness-typical symptoms have been noted.^1^ Outpatient symptom complexes may differ from the better-known inpatient set. A data-driven set of clinical indicators for COVID-19 would help to identify outpatient symptoms and those who most benefit from limited testing availability.

## Methods

CART recursive partitioning, based on presence or absence of symptoms, was used to create a decision tree to correctly classify enrollees into laboratory-confirmed (RT-PCR) COVID-19 cases.^3^ Demographic and symptom data from 736 patients, ages 18-87 years tested for presence of SARSCoV2 virus from March 29-April 26, 2020 were included.

Symptoms reported at enrollment -- fever, chills, cough, sore throat, shortness of breath, muscle aches, abdominal pain, nausea/vomiting, diarrhea, headache, decrease or loss of taste or smell and contact with COVID-19 case -- were included in the CART model as categorical predictors. Presence or absence of SARSCoV2 was the target variable. The tree was built on a ≥10% sample for the parent node and a stopping rule of ≥5% of the sample in the terminal node. A tenfold cross-validation method was used to evaluate reliability. To avoid overfitting, a maximum acceptable difference in risk between the pruned and the sub-tree of one standard error was used for tree pruning. Missing data were handled by surrogate splits. Hosmer-Lemeshow goodness of fit test confirmed the suitability of the trees. The area under the curve (AUC) for receiver operating characteristics, sensitivity, specificity, positive and negative predictive values were assessed. The University of Pittsburgh IRB approved this study.

## Results

Of 736 tested, 55 were positive for SARS-CoV2. Those who tested positive significantly more frequently reported chills, loss of taste/smell, diarrhea, fever, nausea/vomiting and contact with a COVID-19 case, but less frequently reported shortness of breath and sore throat (Table). The CART analyses resulted in a 7- terminal node tree with a sensitivity of 96% and specificity of 53%, resulting in an AUC of 78%. The positive predictive value for this tree was 14% while the negative predictive value was 99%. Of clinical utility, three variables (contact with a COVID-19 case, report of loss or diminution of taste or smell and report of nausea or vomiting; followed down the left side of the Figure) would allow almost half (44%) of the participants to be ruled out as likely non-cases *without* testing.

**Table.** Presenting Symptoms for Outpatients Attending the COVID-19 Testing Clinic

| Symptom | COVID-19 cases<br>N=55 (7.5%)<br>n (%) | Non-cases<br>N=681 (92.5%)<br>n (%) | P value |
| --- | --- | --- | --- |
| Abdominal pain | 20% | 27% | 0.286 |
| Chills | 80% | 64% | **0.016** |
| Cough | 85% | 87% | 0.662 |
| Decrease or loss of taste or smell | 72% | 25% | **<0.001** |
| Diarrhea | 53% | 38% | **0.028** |
| Fever | 86% | 68% | **0.006** |
| Headache | 86% | 82% | 0.497 |
| Muscle aches | 66% | 67% | 0.849 |
| Nausea/Vomiting | 20% | 10% | **0.013** |
| Shortness of breath | 53% | 66% | **0.046** |
| Sore throat | 39% | 66% | **<.001** |
| Contact with COVID-19 case | 70% | 21% | **<.001** |

**Figure.**
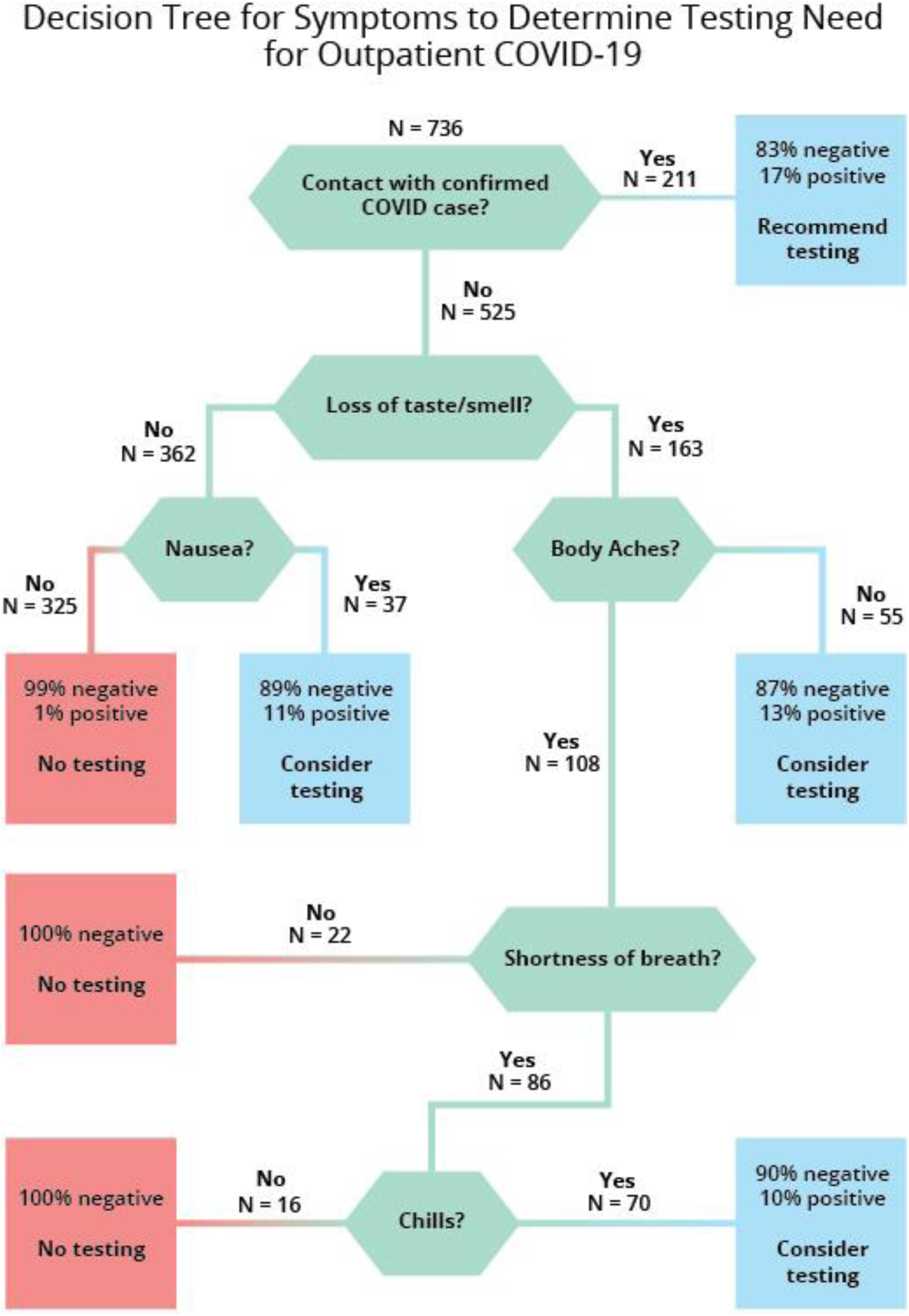

## Discussion

Among those referred for testing, we found negative responses to three questions could classify about half of tested persons who had a minute risk for SARS-CoV2 and would save limited testing resources. These questions are: was the patient in contact with a COVID-19 case? Has the patient experienced 1) a loss of taste or smell; or 2) nausea or vomiting? If COVID-19 case exposure did not occur, and the patient reported neither loss of taste or smell nor nausea or vomiting the patient need not be tested. Alternatively, a clinical decision rule can be electronically programmed to make use of such data. Finally, the outpatient symptoms of COVID-19 appear to be broader than the well-known inpatient syndrome.

## Data Availability

Data will be made available upon request after publication.

## Acknowledgements/Potential Conflict of Interest

Drs. Zimmerman, Nowalk, and Balasubramani and Ms. Eng and Ms. Sax have research funding from Merck & Co, Inc. for unrelated projects. Dr. Zimmerman has funding from Sanofi for an unrelated project. Drs. Bear and Taber have no conflicts to report.

